# Development of a Seizure Matching System for Clinical Decision Making in Epilepsy Surgery

**DOI:** 10.1101/2024.01.21.24301546

**Authors:** John Thomas, Chifaou Abdallah, Kassem Jaber, Olivier Aron, Irena Doležalová, Vadym Gnatkovsky, Daniel Mansilla, Päivi Nevalainen, Raluca Pana, Stephan Schuele, Jaysingh Singh, Ana Suller-Marti, Alexandra Urban, Jeffery Hall, François Dubeau, Jean Gotman, Birgit Frauscher

## Abstract

**Background and Objectives:** The proportion of patients becoming seizure-free after epilepsy surgery has stagnated. Large multi-center stereo-electroencephalography datasets can potentially allow comparing a new patient to past similar cases and make clinical decisions with the knowledge of how similar cases were treated in the past. However, the complexity of these evaluations makes the manual search for similar patients in a large database impractical. We aim to develop an automated system that electrographically and anatomically matches seizures from a patient to those in a database. In addition, since we do not know what features define seizure similarity, particularly considering the various stereo-electroencephalography implantation schemes, we evaluate the agreement and features among experts in classifying seizure similarity.

**Methods:** We utilized SEEG seizures from consecutive patients who underwent stereo-electroencephalography for epilepsy surgery. Eight international experts evaluated seizure-pair similarity using a four-level similarity score through a graphical user interface. As our primary outcome, we developed and validated an automated seizure matching system by employing a leave-one-expert-out approach. Secondary outcomes included the inter-rater agreement and features for classifying seizure similarity.

**Results:** 320 SEEG seizures from 95 patients were utilized. The seizure matching system achieved an area-under-the-curve of 0.82 (95% CI, 0.819-0.822), indicating its feasibility. Six distinct seizure similarity features were identified and proved effective: onset region, onset pattern, propagation region, duration, extent of spread, and propagation speed. Among these features, the onset region showed the strongest correlation with expert scores (Spearman’s rho=0.75, *p*<0.001). Additionally, the moderate inter-rater agreement confirmed the practicality of our approach: for the four-level classification, median agreement was 73.9% (interquartile range, 7%), and beyond-chance Gwet’s kappa was 0.45 (0.16); for the binary classification of similar vs. not related, agreement stood at 71.9% (4.7%) with a kappa of 0.46 (0.13).

**Discussion:** We demonstrate the feasibility and validity of a stereo-electroencephalography seizure matching system across patients, effectively mirroring the expertise of epileptologists. This novel system can identify patients with seizures similar to that of a patient being evaluated, thus optimizing the treatment plan by considering the treatment and the results of treating similar patients in the past, potentially resulting in an improved surgery outcome.

## Introduction

Epilepsy is a prevalent neurological disorder affecting approximately 70 million people worldwide.^1^ About 40% of patients have drug-resistant epilepsy, i.e., they do not respond to anti-seizure medications.^2^ The main potential cure for these patients is surgery. Stereo-EEG (SEEG) has gained acceptance around the world,^3^ as precise localization of the epileptic focus at the sublobar-level is necessary to achieve seizure freedom with minimal functional deficits. Despite these advances, the success rate of epilepsy surgery has been suboptimal^2^ and has stagnated over the past decades. The number of SEEG patients at each center is limited by the complexity of the procedure and there is significant variability in characteristics and implantation strategies across patients.^4–6^ Consequently, most epilepsy centers do not have enough patient cases for epileptologists to optimize the surgery for new patients by comparing with past patients having similar characteristics. Almost every patient presents a novel challenge.

An unexplored avenue for bridging this gap is developing an automated system that can retrieve past patient cases that closely resemble a new incoming patient. Given the individualized nature of epilepsy surgery, such a system could be beneficial for optimizing treatment options. This is particularly important as epilepsy surgery is a form of precision medicine that is individually tailored to each patient. Large open SEEG datasets are now available^7–9^ and others with several thousand patients could be developed in the future. An automated system may have the potential to aid epileptologists to review large SEEG databases rapidly, learn from past patients, and optimize treatment plans of new patients,^10^ something that is impractical to do manually. The identification of electrographically and anatomically similar seizures is the first step towards matching patients. However, what makes experts consider SEEG seizures similar is not clear, particularly in the context of different electrode positions. Hence, developing an automated seizure matching system requires a quantitative study of expert inter-rater agreement (IRA) regarding seizure similarity and the definition of which features are relevant for experts in assessing seizure similarity. Several studies report that the inter-rater agreement for epileptic EEG patterns such as interictal epileptiform discharges,^11^ ictal-interictal-continuum,^12^ neonatal seizures,^13^ and intracranial EEG high frequency oscillations,^14^ may be low to moderate. However, similar studies on pre-surgical planning in epilepsy surgery remain limited.^15–17^

We assessed the expert IRA in determining if SEEG seizures gathered from different patients are indeed similar and which features contributed to this similarity. Based on evidence from scalp EEG,^11,12^ we hypothesized that there exists a moderate agreement for seizure similarity across epileptologists and that a generalizable automated seizure matching system can therefore be developed. First, we measured the IRA across experts in considering two seizures similar by providing information such as the SEEG seizure signals, implantation scheme, and the anatomical regions corresponding to the implanted electrodes, similar to a typical clinical setting. Second, using the similarity scores provided by the experts, we investigated the features considered relevant in matching seizures. Finally, utilizing these features, we propose a reliable automated seizure matching system for improving pre-surgical planning in epilepsy.

## Materials and methods

### Study Design and Patient Selection

Ninety-five consecutive patients who underwent SEEG with imaging at the Montreal Neurological Institute and Hospital (MNI) between 2009-2019 were analyzed (Supplementary Table 1). If a patient had multiple surgeries, we considered the SEEG before the last surgery. We considered all the patients with complete neuroimaging datasets who had at least one artifact-free electroclinical seizure recorded after 48 hours post-implantation. The recordings were acquired with either Harmonie (Stellate) or Nihon-Kohden amplifiers, using homemade MNI or commercial DIXI electrodes.^18^ This study was approved by the MNI Ethics Board (IRB 2019-4880). Written informed consent was obtained from all patients. Our primary objective was to develop an automated system for matching seizures. However, as we do not know what features define seizure similarity, particularly considering the various stereo-electroencephalography implantation schemes, we also included the crucial step to evaluate the agreement among experts in classifying seizure similarity. We conducted the interrater agreement study following the Standards for Reporting Diagnostic Accuracy (STARD) guidelines.^19^ The independent seizure similarity marking by experts who had at least one year of SEEG training for epilepsy surgery serves as index test. The reference standard is the consensus (majority agreement) in seizure similarity among these experts. The study was conducted in three phases (Figure 1A).

**Figure 1:**
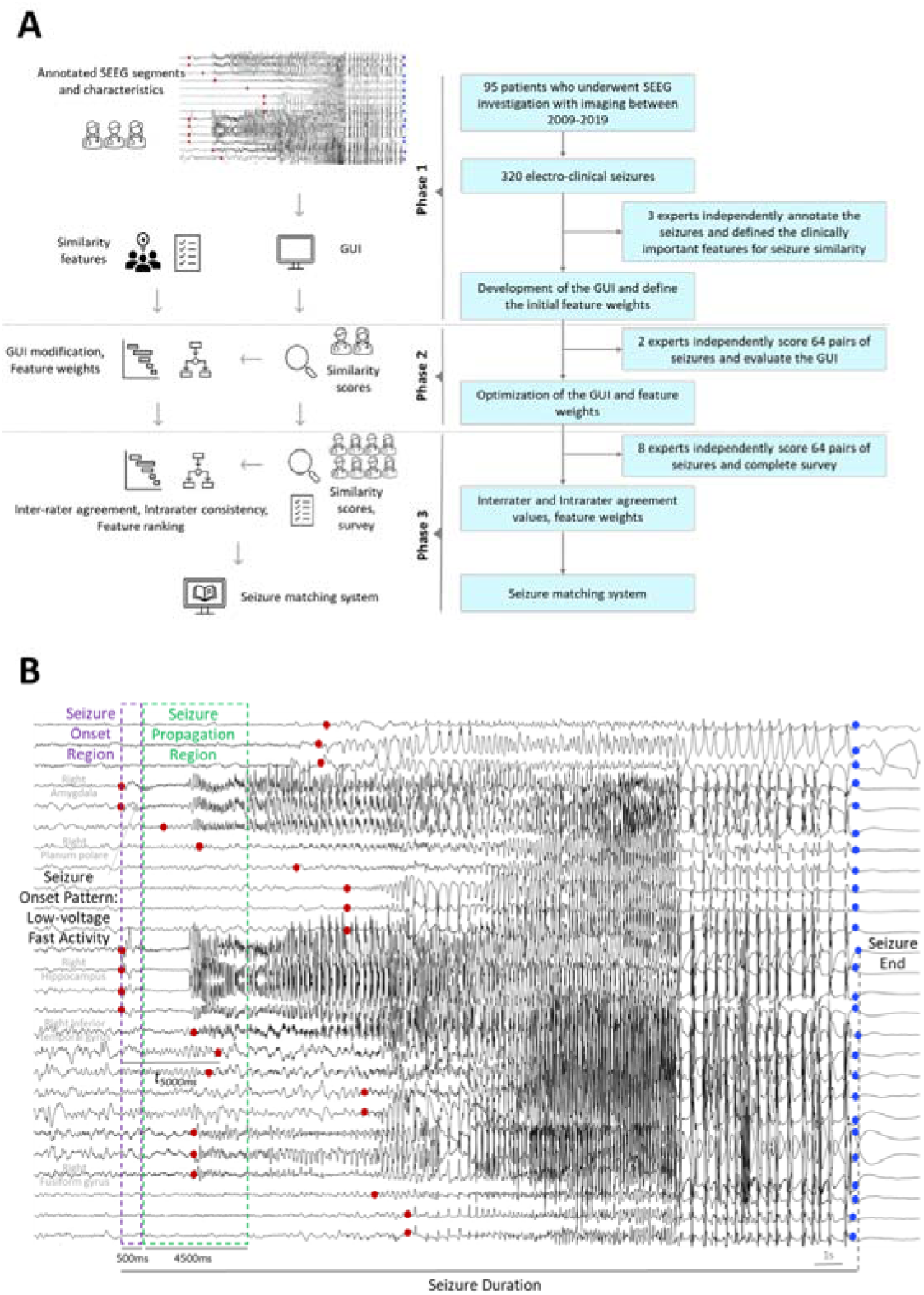
Illustration of study pipeline and seizure similarity features (A) Flow diagram illustrating the study in three phases. Phase 1 involved seizure selection, seizure marking, graphical user interface (GUI) development, and identification of seizure similarity features. Phase 2 included GUI and feature evaluation by two independent experts, and Phase 3 encompassed global evaluation of seizure similarity by eight independent experts. **(B)** Illustration of the six features utilized in this study on a seizure from patient ID 38 with right mesiotemporal lobe epilepsy. In this example, the seizure onset includes six contacts (purple box), while the seizure propagation region includes eight contacts (green box). The seizure onset region was in the right hippocampus and amygdala (atlas regions) and the seizure propagation region included the right fusiform gyrus, planum polare, and the inferior temporal gyrus. Seizure spread is determined as the mean distance covered by the seizure within a 5-second interval, in this case 14.9 mm; seizure speed is measured as the maximum distance covered by the seizure within the same 5-second interval (specifically at t5000 ms), in this case 6.4 mm/s. The seizure duration was 27.2 seconds, and the seizure onset pattern is classified as low voltage fast activity (LVFA).

#### Phase 1: Seizure characterization

A total of 320 electro-clinical seizures were extracted. To avoid overrepresentation of any patient, we selected a maximum of three seizures per seizure-type per patient. The seizure characteristics were considered independent of vigilance state.^20^ For each seizure, the onset, the end, and the onset pattern^21^ were marked on each channel by two board certified neurophysiologists. When the two experts disagreed, a third neurophysiologist was consulted to reach a consensus. The onset was marked as the first sustained rhythmic change in SEEG from the background lasting at least ten seconds independent of the presence of fast activity.^22^ We mapped each SEEG channel into anatomical regions using the 140-region MICCAI atlas.^23,24^

To compare pairs of seizures from different patients, we used a graphical user interface (GUI, Supplementary Figure 1) that integrated multimodal data including implantation scheme, SEEG signals, and the expert markings mentioned above; the comparison resulted in a four-level similarity score (very similar, somewhat similar, low similarity, not related). Next, through a collaborative process involving in-depth discussions with the clinical epileptologists who marked the SEEG, we identified six clinically relevant features for determining seizure similarity: onset region(s), propagation region(s), duration, spread, propagation speed, and onset pattern^21^ (Figure 1B). The detailed definitions and implementation of these features are given in Supplementary S1. An example seizure pair utilized in this study is illustrated in Figure 2. Lastly, we used the following initial weights for the features after consulting with the three experts: 0.5 for onset region and 0.1 for each remaining feature. With these weights (w_l_) for the six feature values (f_l_), we computed the initial similarities,

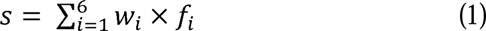

**Figure 2:**
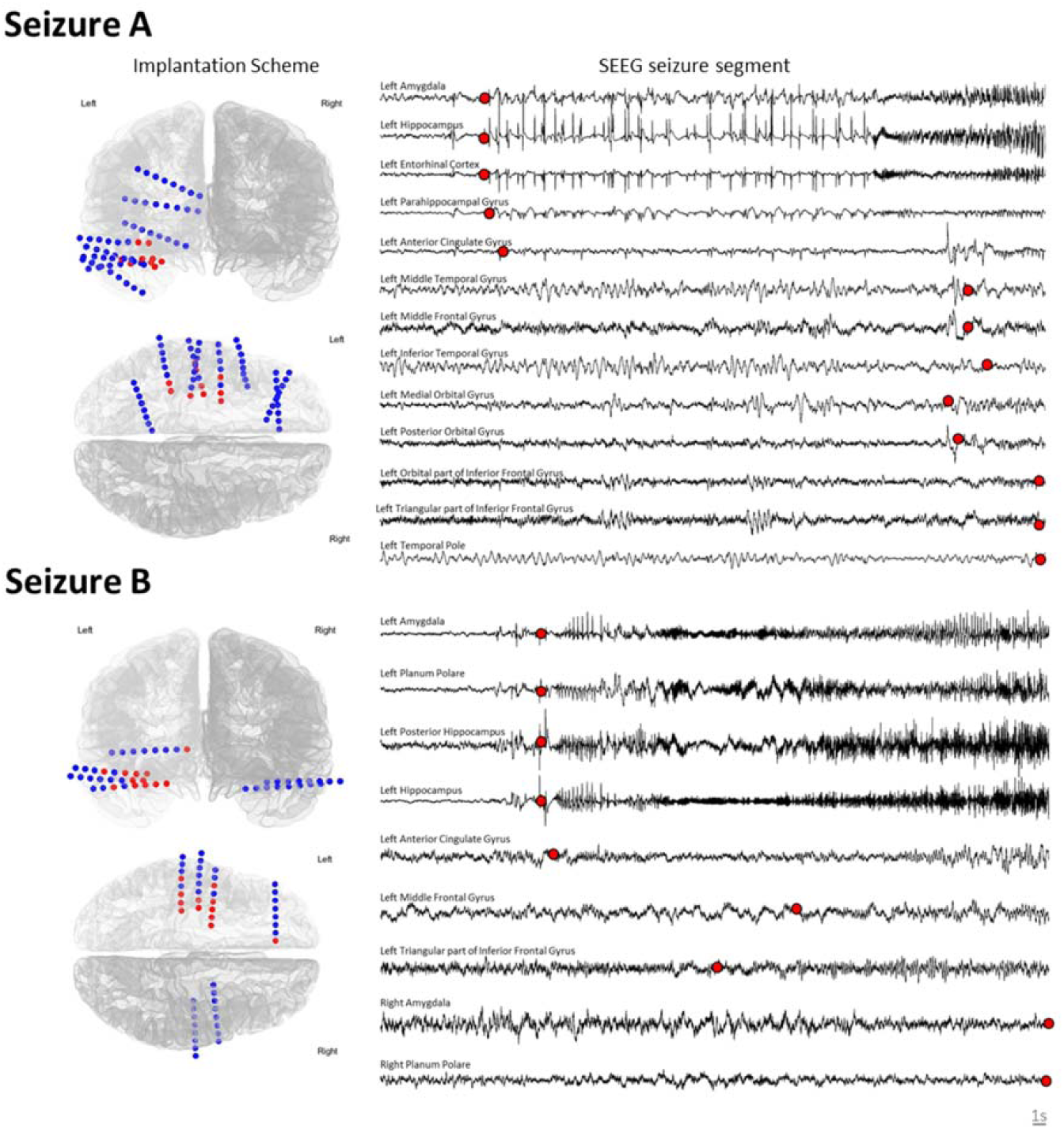
An example of a seizure pair utilized in this study. We illustrate axial and coronal views of the implantation scheme, a 50 second (>ten second pre-ictal) SEEG seizure segment, the brain anatomical locations corresponding to the bipolar channels, and the seizure onset marking on each channel. In the implantation scheme (left), each circle represents a bipolar channel, an array of circles represents an SEEG electrode, and the red channels indicate the bipolar channels involved in the seizure at one-second after the earliest onset of the seizure. In the SEEG seizure segment (right), we illustrate signals filtered between 0.3-100 Hz. Each red circle indicates the onset of the seizure on the particular bipolar channel. Seizure A (patient A) had eight left electrodes targeting mesio-temporal, cingulate, and frontal regions, and seizure B (patient B) had a bilateral implantation with four left and two right electrodes targeting bilateral mesial temporal structures and the left frontal region. We employed six clinically relevant features to assess the similarity between seizure pairs: seizure onset region, propagation region, duration, spread, propagation speed, and onset pattern. Both seizures exhibited a quasi-similar onset in the left mesial temporal structures. In seizure A, the onset was observed in the left amygdala, hippocampus, entorhinal cortex, and parahippocampal gyrus. On the other hand, seizure B showed an onset in the left amygdala, hippocampus, mesial part of the planum polare, and the posterior hippocampus. Both seizures showed early propagation to the mesial frontal area (anterior cingulate cortex) and an onset pattern characterized by spike-wave followed by low voltage fast activity. Seizure A and B lasted for a similar duration of 86.4 seconds and 103.4 seconds, respectively. The spread was consistent at 42.5mm for both seizures, while propagation speeds differed: 66 mm/s for seizure A and 100.7 mm/s for seizure B. The similarity values ranging between 0 and 1 (with 1 indicating high similarity) for each feature between this seizure pair were as follows: onset region (f_l_, 0.95), propagation region (f_2_, 0.91), duration (f_3_, 0.84), spread (f_4_, 0.99), propagation speed (f_5_, 0.65), and onset pattern (f_6_, 1). This seizure pair was marked as similar by seven experts (‘Very similar’ by five, and ‘Somewhat similar’ by two experts). Detailed definitions and implementation of these features are given in Supplementary S2.

#### Phase 2: Preliminary assessment of GUI and features

Two independent experts not involved in phase 1 evaluated the GUI and scoring of seizure similarity. Each expert assessed 64 pairs of seizures from different patients, randomly selected according to the initial similarity scores, with 16 (25%) allocated to each similarity-level. The experts provided the following suggestions regarding the interface: (1) incorporate two sagittal (left and right) and coronal views of the implantation scheme, along with an axial view. (2) include anatomical locations corresponding to each SEEG channel. (3) implement an SEEG segment that allows navigation back and forth for at least 100 seconds, with a visible time indicator. The GUI was modified according to their suggestions. The feature weights were adjusted to 0.4 for onset region, 0.2 for propagation region, and 0.1 each for the remaining features, by correlating the feature values with expert scores (Spearman’s rho), and the new similarities were computed.

#### Phase 3: Evaluation of seizure similarity

To ensure the generalizability of the results and mitigate potential bias from similar training and expertise, we selected eight experts trained at different tertiary epilepsy centers around the world, who did not participate in phases 1 and 2. Each expert assessed 64 pairs of seizures each, with 16 (25%) allocated a priori to each similarity-level. Similarity levels were defined with the weights initially imposed for each feature to ensure a well-balanced distribution of seizure pairs for the experts (some very similar, some very different…). These weights did not influence their evaluation of features. The features were not disclosed to the experts apriori, allowing them the autonomy to assign similarity scores based on their training and expertise. Thirty-two pairs were identical across the experts. Twenty-two pairs were randomly selected, and the final ten pairs were flipped pairs randomly chosen from the set of 54 (32+22). Based on these scores, we investigated the IRA, intra-rater consistency, univariate/multivariate feature assessment, and developed the seizure matching system.. Further, we conducted a survey among experts to analyze their opinions on the seizure matching system and the relevant features for matching (Supplementary S2).

The three analyses, namely, the univariate, multivariate, and survey-based, provided three distinct pieces of information. The univariate analysis gave us an estimate whether the selected features were important for seizure similarity. The multivariate results provided insights into the optimal combination of features for determining similarity, considering correlations between features. Lastly, survey-based opinions offered an understanding of how experts perceived the relative importance of the features.

### Evaluation and Statistical Analysis

#### Intra-rater consistency and Inter-rater agreement

We quantified the intra-rater consistency for each expert by calculating the percentage agreement between the similarity scores of the ten flipped seizure pairs. We applied the Stuart-Maxwell’s test for marginal homogeneity to examine whether experts categorized seizure similarity into four-levels with comparable proportions.^25^ The p-values were corrected for multiple comparisons by controlling for the false discovery rate.^26^

The pairwise IRA was determined based on the 32 identical seizure pair similarity scores, by calculating the percentage agreement (P) and chance adjusted Gwet’s kappa.^27,28^ Kappa was calculated as (P_a_ - P_C_)/(1 - P_C_), where P_C_ is the chance agreement. Kappa was computed in two scenarios: considering the four-level similarity scores, and by combining the scores into binary labels: similar (including “very similar” and “somewhat similar”), vs. not related (including “less similarity” and “not related”). We utilized standard convention for interpreting of kappa; 0-0.20 (slight), 0.21-0.40 (fair), 0.41-0.60 (moderate), 0.61-0.80 (substantial), and 0.81-1.00 (almost perfect).^29^

We also compared the predicted probability of an expert assigning a seizure pair as similar (binary labels) to the group event probability by applying statistical calibration measures. This shows how well the expert prediction probabilities agree with the group probability. First, we binned the seizure pairs into five bins (similarity deemed by one expert, similarity deemed by two or three experts, similarity deemed by four or five experts, similarity deemed by six or seven experts, and similarity deemed by all eight experts). We opted for five bins to maintain a reasonable number of data points in each bin, similar to what has been adopted in the literature^12^. Next, each expert’s prediction probability was defined for each bin as the proportion of seizure pairs in the bin that were marked as similar by the expert. For each expert, the calibration curve is estimated with the group probabilities on the x-axis and the expert’s prediction probability on the y-axis. A calibration score per expert was also defined as the mean absolute value of the difference between an individual’s prediction probabilities and the group probabilities. Significant overcalling and undercalling were defined as having a calibration score above 0.2.^12^ A calibration score lower than 0.2 indicated that the expert predictions were in alignment with the group probability.

#### Seizure matching system and analysis of features

All 64 seizure pairs were considered for univariate/multivariate feature assessment and the development of the seizure matching system. Here, we transformed the similarity scores into numerical values using a uniform scale (0, 0.33, 0.66, and 1), representing a spectrum from ‘not related’ to ‘very similar’. In univariate analysis, we investigated how well each feature correlated with the individual and mean expert scores utilizing Spearman rho. To test that the correlation values were significant, we determined p-values by generating a distribution of correlation values by randomly permuting expert scores. The correlations between the six features were also quantized.

We utilized an ensemble decision tree^30^ implementing the seizure matching system. The parameters of the decision tree are optimized using Bayesian optimization^31^. To validate the model and the features, a leave-one-expert-out (LOEO) cross-validation was employed using the binary labels. In each step, the seizure pairs marked by one expert were withheld as the test set. Using the scores from the remaining experts, a model was trained to predict the labels for the left-out expert. This procedure was repeated for each expert and LOEO validation had eight runs. The performance of the model was evaluated with the area-under-the-receiver-operating-characteristics curve (AUC) and area-under-the-precision-recall curve (AUPRC). To ensure robustness of the results, given that each model training initiates with a random set of hyperparameters, the LOEO was iterated 100 times (eight runsX 100), and the results were averaged.

The feature importance was determined based on Gini’s diversity index in each step of LOEO and was averaged. The feature importance was determined based on Gini’s diversity index in each step of LOEO and was averaged. Decision trees have the capability to identify the most informative features during the training process. The algorithm evaluates various combinations of features at each of the decision tree nodes and chooses the combination that contributes the most to the classification performance, reducing redundancy. Consequently, it takes care of correlated features automatically. For each expert, we also compared the model prediction with the four-level similarity scores using Spearman’s rho. Finally, the survey-based expert opinions regarding the features were compared with the results of the LOEO feature importance to identify potential correlations or discrepancies. The evaluation and analysis was performed using MATLAB 2020^32^ and IRRCAC python toolbox.^28^

### Data Availability

The data that support the findings of this study are available upon reasonable request and if in accordance with the respective Research Ethics Boards’ policies.

## Results

A total of 320 seizures from 95 patients were analyzed in this study. The patient cohort was diverse: 26 (27.4%) had right, 35 (36.8%) had left, and 34 had bilateral SEEG electrode implantations; 46 of the 95 patients (48.4%) were female with age=32.5 (17, median (interquartile range)). Temporal lobe epilepsy was present in 60% (57 patients). Preoperative magnetic resonance imaging (MRI) revealed mesial temporal sclerosis in seven (7.3%), focal cortical dysplasia in 20 patients (21.0%), no abnormality in 23 (24.2%), and 45 patients (47.4%) exhibited tumors, cystic lesions, traumatic lesions, or other abnormalities.

### Seizure matching system

We utilized an ensemble decision tree to implement the seizure matching system. It achieved an AUC of 0.820 (95% CI, 0.819-0.822) and AUPRC=0.723 (95% CI, 0.719-0.727) for the LOEO validation (across 100 iterations). The AUC was also consistent across experts (range 0.704-0.907, Figure 3). Further, the seizure matching system predictions for each expert were significantly correlated (each *p*<0.01) with the four-level similarity scores with rho=0.54 (0.21).

**Figure 3:**
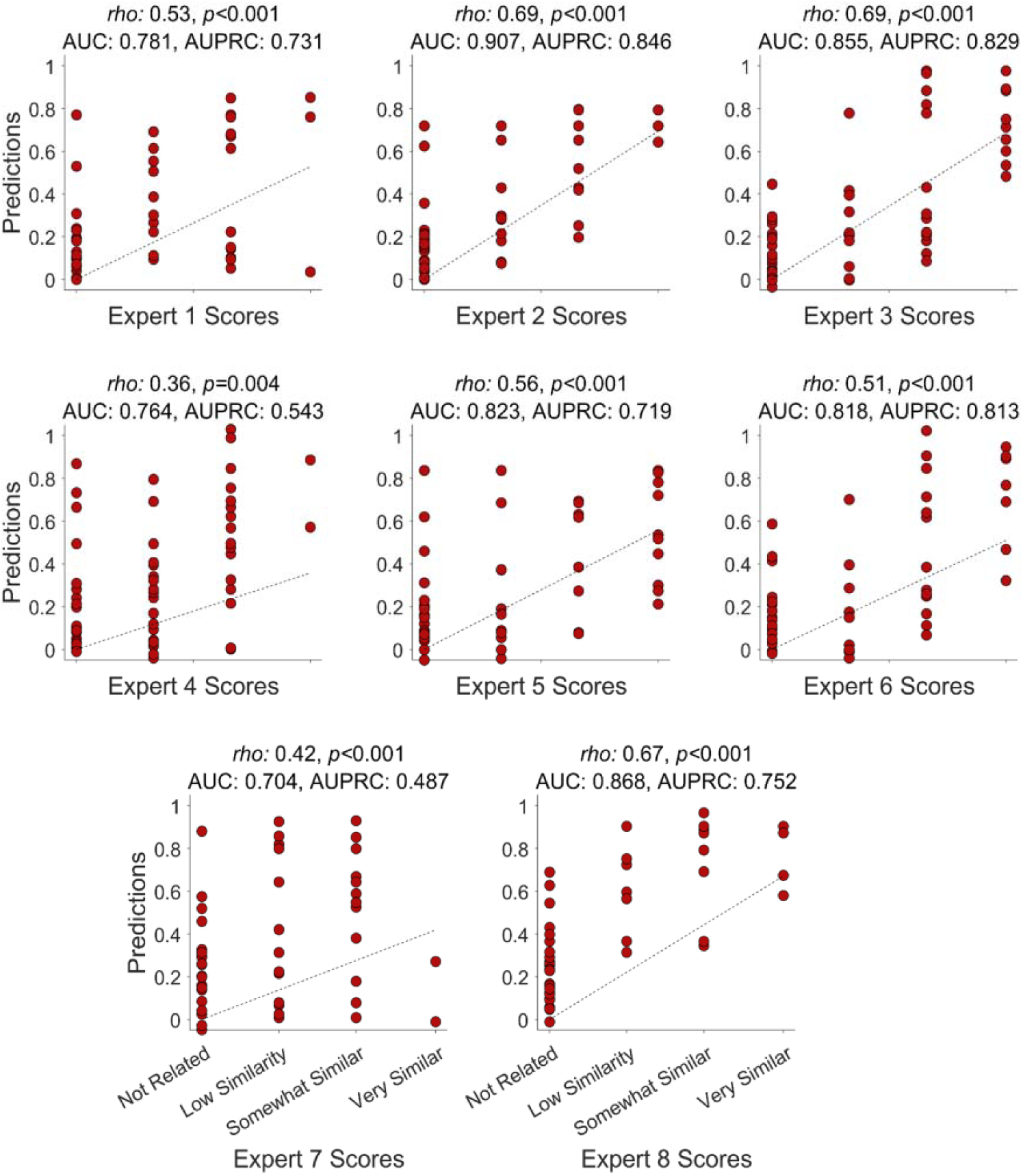
Leave-one-expert-out results for each expert. Here we illustrate the predictions in each run of the leave-one-expert-out cross-validation (y-axis) in comparison with the labels from the left-out expert (x-axis). Each red circle represents the prediction value of similarity across a pair of seizures. The prediction values ranged from 0 and 1, with a higher value indicating better similarity between the seizures. The rho and p-values are computed for the prediction values vs. the four-level similarity scores. The rho values were statistically significant for all the experts. In addition, we have also provided the area-under-curve and area-under-precision-recall curve are computed based on binary labels (similar vs. not related) for each run.

### Intra-rater consistency and Interrater agreement

We developed a multimodal GUI to compare similarity across pairs of seizures and modified it based on phase 2 comments from two experts. Subsequently, we utilized it to evaluate seizure similarity scores from eight global experts. Utilizing the similarity scores from the experts, we evaluated the intra-rater consistency and the IRA. The intra-rater consistency was high at 100% (25%) (median (interquartile range), four-level classification), and 100% (0%), allowing for a single-level error, for example, categorizing the original pair as ‘low similarity’ and categorizing the same pair as ‘not related’ when flipped. The IRA was moderate: four-level classification: agreement=73.9% (7%), beyond-chance agreement Gwet’s kappa=0.45 (0.16); binary classification of similar vs. not related, agreement=71.9% (4.7%), kappa=0.46 (0.13); Figure 4A, B. The experts grouped seizure similarity comparably; the Stuart-Maxwell test achieved non-significant results (*p*>0.05) for all combinations of experts. The mean calibration scores for all experts were 0.11 (0.05), lower than 0.2, indicating that their marking aligned with the group probability (Figure 4C).

**Figure 4:**
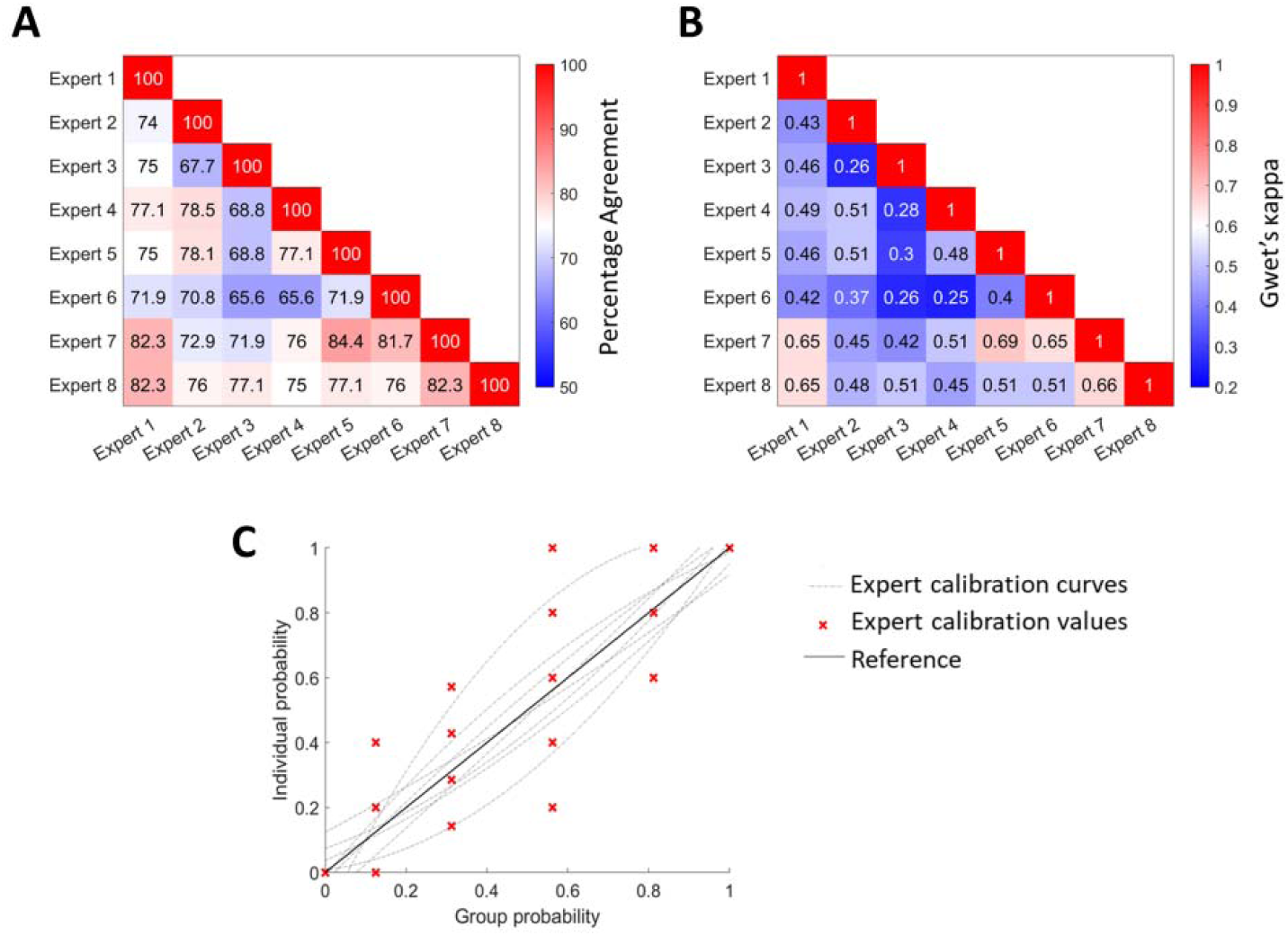
Inter-rater comparison across eight experts for seizure similarity: **(A)** four-level classification percentage agreement, **(B)** four-level classification beyond-chance Gwet’s kappa. The percentage agreement and kappa were moderate implying there exists reasonable agreement in seizure similarity across experts. **(C)** Calibration curves for the eight experts for binary similarity scores with group probability in five bins. For each individual expert, we applied polynomial fitting of degree two to estimate the calibration curve. The individual expert values are represented in red. Most of the individual calibration curves align with the reference line (black). For each group probability bin, the red markers indicate an expert individual probability which may overlap with other experts.

### Analysis of features

We determined the feature ranking for seizure similarity using univariate and multivariate analysis. Out of the six features, only the onset region and the propagation region were correlated with each other (rho=0.81, *p*<0.001, Supplementary Figure 3). The onset region was identified as the most correlated feature with the expert scores: univariate analysis with mean expert scores rho=0.75, *p*<0.001, Figure 5; univariate analysis with individual expert scores rho=0.52 (0.17), each *p*<0.05, Figure 6A; multivariate LOEO feature importance=0.79 (0.09), Figure 6B.

**Figure 5:**
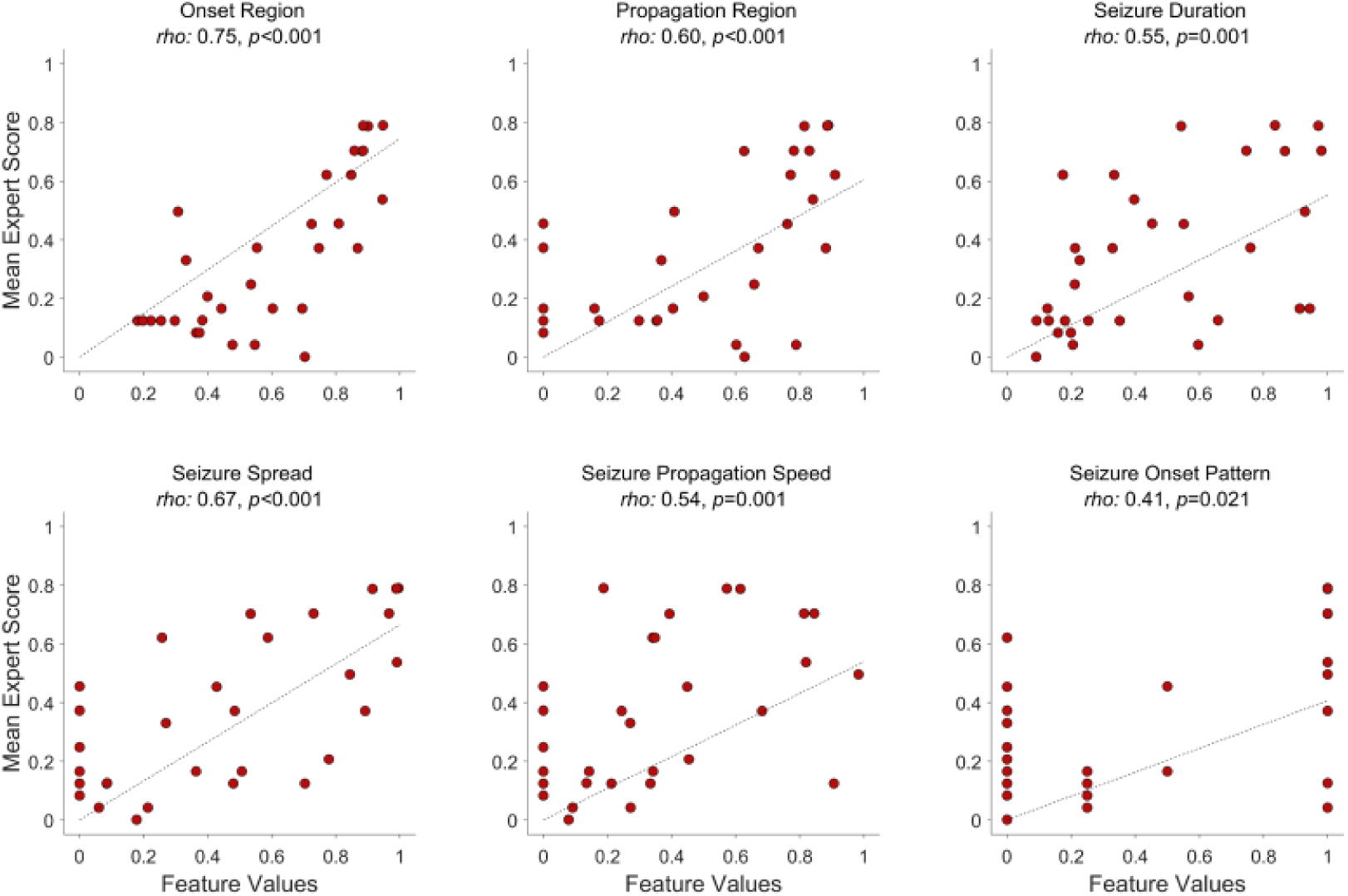
Univariate analysis of features for seizure similarity with the mean expert similarity scores. Here we illustrate the mean expert scores for each pair of seizures (y-axis) vs. the feature values (x-axis). The expert scores ranged from 0 and 1, with a higher value indicating better similarity between the seizures. Each red circle represents the similarity score of a pair of seizures. In addition, we also illustrate the correlation line that has been fitted through these points. Rho value ranges between [0, 1] with a high rho value indicating better correlation. The rho values were moderate to high for the six features. We used subjective correlation values for evaluating the similarity across seizure onset patterns based on the opinion of experts (Supplementary Figure 1). The seizure onset region had the highest rho value with the mean expert scores.

**Figure 6:**
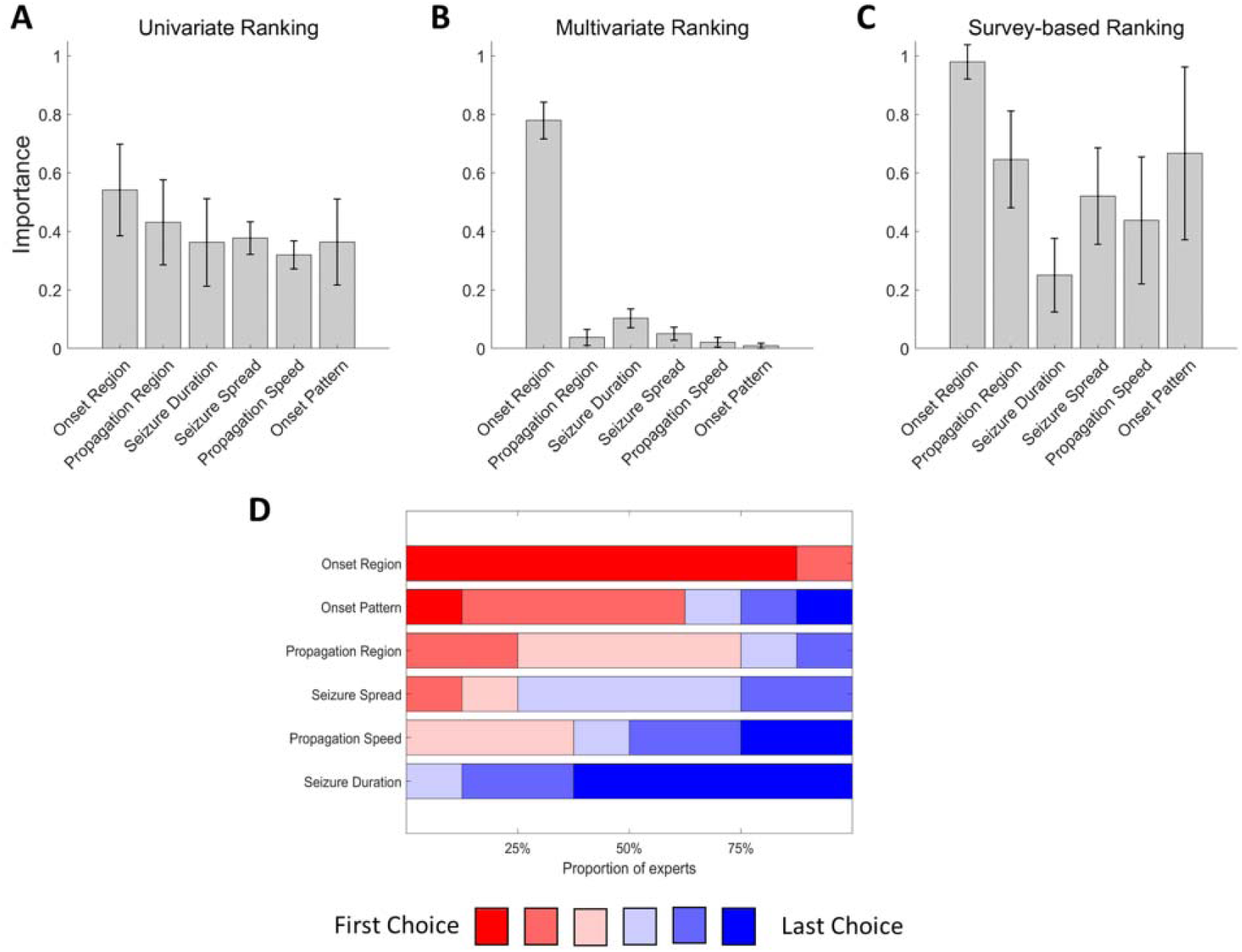
Seizure similarity feature ranking determined by univariate, multivariate, and survey-based analysis. **(A)** Spearman rho values derived for each feature across the eight expert scores. Overall, the onset region had higher mean rho values indicating higher correlation with the expert scores. **(B)** Feature importance derived based on leave-one-expert-out (LOEO) results for binary similarity classification. The onset region was identified as the most important feature to determine seizure similarity. **(C)** Feature importance derived based on expert opinion survey. The experts also suggested the onset region as the most important feature in determining seizure similarity. **(D)** Detailed results of feature importance of the expert opinion survey. The onset region was ranked the best feature (seven out of eight experts).

### Survey results

All eight experts unanimously agreed on the benefits of developing a seizure matching tool. Additionally, they rated the GUI with a mean score of 8.6/10 for its utility. The onset region was ranked as the most important and the seizure duration the least important feature by the experts (Figure 6C, D). Most of the experts (6/8) considered the seizure characteristics independent of the hemisphere. Four experts were satisfied with the features considered in this study. Two experts recommended utilizing seizure semiology, and one expert emphasized the use of the underlying etiology of epilepsy.

## Discussion

Despite many advances in SEEG analysis over the past decades, results of surgical outcome have not clearly improved. One potential reason may be a lack of systems allowing matching of past patients to patients currently investigated, and benefitting from the experience gained from the past. In this study, we sought to address the issue by developing a generalizable automated seizure matching system. Prior to developing an automated system, we had to determine if there was sufficient agreement among experts when it comes to assessing if two seizures, from different patients with different implantation schemes, can be considered similar. The three main findings of this study are: (i) the automated seizure matching system, evaluated by LOEO cross-validation, yielded an AUC=0.82, demonstrating the feasibility of developing a reliable clinical system; (ii) there exists a moderate agreement across experts in determining similarity across SEEG seizures from different patients with different electrode locations; (ii) the mean expert similarity scores were correlated with six clinically identified features, with the onset region being the highest ranked feature.

### A step towards a reliable patient matching system

In the present investigation, we focused on developing a seizure matching system aimed at assisting epileptologists in planning epilepsy surgery with the help of the analysis of past similar patients. The existing literature primarily concentrates on optimizing the pre-surgical evaluation of epilepsy using methods or biomarkers for identification of the epileptogenic zone^7,33,34^ or prognostication of surgery outcomes.^35–38^ Given that we have achieved a moderate IRA (kappa=0.45) and a satisfactory AUC=0.82, we were successful in demonstrating the feasibility of a seizure matching system. This system can be readily integrated into current clinical workflows. Its usefulness will increase with the size of the database available, and it will be optimal with a multicenter database much larger than what is usually available in one center. For example, when a new patient presents with similar seizures to that of a database patient, the physician can evaluate the type and extent of resection that were performed, the outcome, and determine if this can positively impact the treatment of the current patient. This is a unique approach in a way that it allows the physician to learn from a large number of prior patient cases.

### High intra-rater consistency and moderate inter-rater agreement

The intra-rater consistency in judging seizure similarity was high, indicating consistent scores among the experts. Moreover, the experts categorized seizure similarity into four-levels with a comparable marginal probability. Many studies^11,12^ failed to assess intra-rater consistency prior to evaluating inter-rater agreement. Moderate IRA among experts (kappa=0.45) was observed in assessing seizure similarity across a sizable, diverse set of patients. This corroborated with the IRA related to interictal and ictal scalp EEG findings ranging between 0.39-0.48.^11–13^ There are two main studies that have investigated agreement across aspects of pre-surgical planning of epilepsy using invasive EEG: seizure localization (excellent agreement, intra-class correlation coefficient=0.79, seven centers, number of experts not reported)^16^ and seizure-onset zone identification (substantial, kappa=0.8, two experts).^17^ However, both of these studies were performed on a small number of patients (10 and 11) and almost exclusively with temporal lobe epilepsy. It must also be noted that the task itself, matching seizures from patients with different implantation schemes, is new to epileptologists, probably explaining some of the diversity in the approaches taken by the experts in this study. We did not observe any undercallers or overcallers among the experts even though they belonged to different institutions and had undergone training in various centers. These findings underscore the feasibility of a generalizable seizure matching system.

### Expert similarity scores correlate with six clinically identified features

The correlation coefficients were statistically significant and moderate to high for all the six features in comparison with the mean expert scores. These findings are consistent with the opinion of epileptologists and suggest that these features hold clinical significance in determining seizure similarity. In contrast to scalp EEG (routine, sleep, or critical care), for which standard guidelines have been established,^39,40^ intracranial EEG lacks international guidelines, and a global standard terminology is yet to be devised. With an AUC=0.82, we were able to satisfactorily quantify the seizure similarity using six features. The seizure onset region exhibited higher significance than all other features across both univariate and multivariate analyses, as well as in survey opinion rankings (Figures 5, 6). This finding is also consistent with the concept of anatomo-electro-clinical correlation, utilized to determine the epileptogenic zone.^41^

Despite aligning with the initial weights assigned based on the opinions of three experts in phase 1 and two experts in phase 2, the autonomy granted to the eight experts in phase 3, along with their diverse training backgrounds, ensured that the feature rankings were not influenced by these initial weights. We currently employed features that appeared to be commonly used in clinical practice. The same approach can be applied to other features that may need to be included in the future. Further, we observed that, in their assessment of feature importance, the experts tended to overestimate the importance of certain features compared to their actual use of these features when they assessed seizure similarity, such as the seizure onset pattern and propagation region. While the onset pattern was ranked as the second-best feature in the survey-based evaluation (Figure 6 C, D), its importance was comparatively lower in the multivariate feature analysis, when considered alongside the onset region (Figure 6B). The onset pattern exhibited no correlation with the onset region (Supplementary Figure 3), suggesting that the onset pattern might not be relevant for patient matching in combination with the onset region. Similar opinions were observed for the propagation region; however, it had a high correlation with the onset region (Supplementary Figure 3).

### Potential avenues for future research and constraints

Seizure similarity serves as the initial step towards achieving patient similarity; it is necessary to consider other factors such as seizure semiology,^42^ MRI findings,^4^ etiology, interictal biomarkers,^35^ implantation scheme, etc., to have a comprehensive understanding of patient similarity and facilitate the phenotyping of patients into groups with shared characteristics, aiding the search for improved personalized treatments. The current study serves a proof-of-principle that a seizure matching system can be implemented in the current clinical context of epilepsy surgery. This personalized approach holds promise for enhancing epilepsy surgeries success rates, leading to improved treatment outcomes and overall patient well-being. This study represents a critical step towards a patient matching system for epilepsy surgery.

In our analysis, we utilized the Euclidean distance to compute the distance between seizure onset regions and between propagation regions. It is crucial to acknowledge that connectivity strength may vary among different brain regions, irrespective of their spatial proximity. We intend to further explore the potential utilization of functional distances to enhance our understanding of seizure similarity. Similarly, we acknowledge the existence of variations in the classification of seizure onset patterns^43^. While we have adhered to a specific classification^21^, a comparable approach can be applied using alternative classifications.

## Conclusions

This study demonstrated the feasibility of an automated system and features for matching seizures across patients, providing a valuable tool to make use of vast numbers of existing cases and complement the expertise of epileptologists in SEEG pre-surgical evaluation. The moderate IRA among experts supports the development of a generalizable automated seizure matching system. The proposed system achieved promising performance, indicating its potential to optimize treatment plans and improve surgical outcomes. It can efficiently review large SEEG databases, aiding in matching similar cases, and facilitating personalized epilepsy surgery investigation and treatment, something that is impractical to achieve manually. Prospective evaluation and collaboration are needed to refine and enhance the system’s capabilities, paving the way for advancements in epilepsy research and improved patient care.

## Acknowledgements

We acknowledge the help and support from Dr. Nicolas von Ellenrieder and the staff and technicians at the EEG Department of the Montreal Neurological Institute and Hospital (Lorraine Allard, Nicole Drouin, and Chantal Lessard).

## Funding

This work was funded by project grants from the Canadian Institutes of Health Research (PJT-175056 to BF, FDN-143208 to JG, and MFE CIHR-IRSC:0633005463 to JT), and Tanenbaum Open Science grant (JT, JG, BF). JT was supported by the Jeanne Timmins Fellowship. BF was supported by a salary award (“Chercheur-boursier clinicien Senior”) of the Fonds de Recherche du Québec – Santé 2021-2023.

## Supplementary material

**Supplementary Table 1.** Clinical patient details

**Supplementary Figure 1:** Graphical User Interface

**Supplementary S1:** Implementation of similarity features across seizure pairs

**Supplementary Figure 2:** Subjective correlation matrix for seizure onset patterns

**Supplementary Figure 3:** Feature correlation matrix

**Supplementary S2:** Survey document

## Notes

### Competing Interest Statement

The authors have declared no competing interest.

### Author Declarations

Ethics Board of Montreal Neurological Institute-Hospital, McGill University, Canada gave ethical approval for this work.

