## Supplementary material for "Development of a Seizure Matching System for Clinical Decision Making in Epilepsy Surgery"

### Supplementary Table 1: Clinical patient details

| **Patient ID** | **Age group** | **Epileptic focus** | **MRI findings** |
| --- | --- | --- | --- |
| 1 | 31-50 | R orbito-frontal, R anterior insular | R orbito-frontal to anterior insula FCD |
| 2 | 18-30 | R posterior cingulate & precuneus | R posterior cingulate FCD |
| 3 | 31-50 | R frontal operculum (face motor area) | R prefrontal cavernoma |
| 4 | <18 | L parietal operculum & supramarginal gyrus | L parietal operculum & supramarginal gyrus FCD |
| 5 | 31-50 | R mesial postcentral gyrus, precuneus, posterior cingulate | R paramedian postcentral gyrus FCD |
| 6 | 31-50 | R temporo-parietal | B hippocampal atrophy, surgical cavity |
| 7 | 18-30 | R posterior cingulate, R precuneus, R anterior & mid-cingulate | R postcingulate lesion |
| 8 | 31-50 | L superior temporal gyrus | Surgical cavity, encephalomalacia |
| 9 | 31-50 | R temporo-parieto-occipital | Normal |
| 10 | 18-30 | L temporo-insular (posterior) | L posterior insula/Heschl gyrus FCD & surgical cavity |
| 11 | 18-30 | L latero-mesial occipital | L latero-occipital FCD |
| 12 | 31-50 | L temporo-occipital | Normal |
| 13 | 31-50 | L temporo-parieto-occipital | L post-quadrant dysplasia, max temporo-occipital (hemi-megalencephaly) |
| 14 | >50 | L mesio & lateral temporal | Normal |
| 15 | 31-50 | L orbito-frontal | Questionable L mesial orbitofrontal blurring suggestive of FCD |
| 16 | 31-50 | L mesio-temporal | L para hippocampal cystic lesion |
| 17 | 18-30 | R fronto-temporal (widespread: orbitofrontal, anterior cingulate, anterior insula, pole frontal & mesio-temporal ) | Normal |
| 18 | 18-30 | R precuneus, R mesial postcentral gyrus, R superior parietal lobule | R precuneus FCD |
| 19 | >50 | R mesio-temporal | R Hippocampal atrophy |
| 20 | 31-50 | R middle frontal gyrus | R mid frontal convexity FCD |
| 21 | 31-50 | B mesio-temporal (Amygdala, Hippocampus) | R frontal encephalomalacia, B hippocampal atrophy |
| 22 | 31-50 | L anterior cingulate, L orbito-frontal, L amygdala | L anterior cingulate FCD |
| 23 | >50 | R insular & R central operculum | Normal |
| 24 | 31-50 | L mesio-temporal | L Hippocampal atrophy |
| 25 | 31-50 | L mesio-temporal | B frontal PNH |
| 26 | 31-50 | L mesio-temporal | L PCA territory remote ischemic lesion |
| 27 | 18-30 | R temporo-insular | Normal |
| 28 | 31-50 | R temporo-insulo-parietal | Diffuse atrophy |
| 29 | 18-30 | L middle frontal gyrus | L precentral, deep (F1 & F2) FCD |
| 30 | 18-30 | L fronto-temporal (orbito-frontal, anterior cingulate, mesio-lateral temporal) | L anterior cingulate/orbito-frontal FCD |
| 31 | 18-30 | L temporo-parietal | B band heterotopia |
| 32 | 18-30 | L hippocampus, posterior cingulate | L MTS & surgical cavity |
| 33 | >50 | R mesial & lateral temporal | Normal |
| 34 | 31-50 | L middle frontal gyrus | Normal |
| 35 | 31-50 | L mesio-temporal | L TMS |
| 36 | 18-30 | R SMA, R mid-cingulate & R mesial superior frontal gyrus | R (F1 & F2) FCD |
| 37 | 18-30 | L & R mesio-temporal | R hippocampal atrophy |
| 38 | 18-30 | R mesio-temporal | R hippocampal hypersignal, L frontal cystic-like lesion |
| 39 | 31-50 | R fronto-temporal | Normal |
| 40 | 18-30 | L mesio-temporal | Normal |
| 41 | 18-30 | R postero-lateral temporal | R frontal atrophy & surgical cavity |
| 42 | 18-30 | L orbito-frontal | Fibrous dysplasia L sphenoid, L orbitofrontal encephalocele (post-op), & surgical cavity |
| 43 | 31-50 | L superior frontal gyrus | L superior frontal gyrus FCD |
| 44 | 31-50 | L mesio-temporal | L hippocampal atrophy |
| 45 | 31-50 | R mesio-temporal | Normal |
| 46 | 18-30 | B temporal | B temporo-occipital PNH |
| 47 | 31-50 | L mesio-temporal | Normal |
| 48 | 18-30 | L fronto-parietal | Normal |
| 49 | 31-50 | B mesio-temporal, R lateral parietal | B parieto-occipital atrophy & gliosis (precuneus & cuneus); surgical cavity |
| 50 | 31-50 | L fronto-parietal | L parietal & insular cyst & gliosis |
| 51 | 31-50 | R fronto-temporal (orbito-frontal, anterior cingulate & mesio-temporal ) | Normal |
| 52 | <18 | R orbito-frontal | R pre-cuneus cystic lesion, R orbito-frontal, anterior cingulate FCD |
| 53 | 18-30 | L posterior insular, parietal operculum | L centro-parietal, post-insular encephalomalacia |
| 54 | 31-50 | L temporo-occipital | L posterior insula, temporal & parietal atrophy & gliosis |
| 55 | 31-50 | R fronto-parieto-temporal | Agenesis of corpus callosum |
| 56 | 31-50 | L temporo-parieto-occipital | L hippocampal atrophy, L fusiform gyrus & pericalcarine atrophy, & mild diffuse L hemispheric atrophy |
| 57 | 18-30 | L mesial & lateral temporal | L mesiotemporal cortico-subcortical blurring |
| 58 | 31-50 | L SMA | Normal |
| 59 | 31-50 | L mesio-temporal (Amygdala, Hippocampus, Parahippocampal gyrus ) | Normal |
| 60 | 18-30 | L temporo-fronto-parietal | Normal |
| 61 | 31-50 | L temporo-fronto-parieto-insular | Normal |
| 62 | 31-50 | L mesio-temporal | Normal |
| 63 | 31-50 | R temporal pole, temporal neocortex, mesiotemporal | PNH in R trigonal area |
| 64 | 31-50 | B mesial & lateral temporal | R hippocampus atrophy |
| 65 | 31-50 | R orbito-frontal | R orbito-frontal FCD |
| 66 | 18-30 | R mesio-temporal | R MTS & L occipital uligyria/ encephalomalacia |
| 67 | 31-50 | R temporo-occipital | R temporo-occipital cyst |
| 68 | 31-50 | L mesio-temporal | L orbito-frontal encephalocele, L hippocampal malrotation |
| 69 | 31-50 | L fronto-insular | Normal |
| 70 | 18-30 | R & L mesio-temporal | Diffuse atrophy |
| 71 | 31-50 | R & L mesio-temporal | Bilateral hippocampal atrophy |
| 72 | >50 | L temporo-insular (posterior) | L Heschl gyrus & posterior insula FCD |
| 73 | 18-30 | L middle frontal gyrus | L frontal (F2) FCD |
| 74 | <18 | R frontal parasagittal | R frontal parasagittal FCD |
| 75 | 18-30 | R orbito-frontal | Normal |
| 76 | 31-50 | R orbito-frontal, R anterior insular, R SMA | R hemi-megalencephaly |
| 77 | >50 | L mesio-temporal | L anterior temporal encephalomalacia; L hippocampal atrophy |
| 78 | 18-30 | R occipito-temporal | Normal |
| 79 | <18 | L mid-cingulate & SMA | L mid-cingulate/inferior SMA FCD (along calloso-marginal sulcus) |
| 80 | 31-50 | L & R mesio-temporal | Normal |
| 81 | 18-30 | R orbito-frontal, R anterior insular, R SMA | Atypical gyrification of R orbito-frontal, R frontal pole |
| 82 | 18-30 | R fusiform gyrus & R posterior nodule | R temporo-occipital nodules, one L occipital nodule |
| 83 | 18-30 | L mesio-temporal | Normal |
| 84 | 31-50 | R insular | Severe B frontal atrophy |
| 85 | 18-30 | L central | Large surgical cavity in L parietal lobe, involved posterior insula |
| 86 | 31-50 | L mesio-temporal | Smaller L hippocampus & unusual gyral formation of L fusiform gyrus |
| 87 | 18-30 | R latero-occipital | Normal |
| 88 | 31-50 | L parieto-occipital | L parieto-occipital lesion |
| 89 | 31-50 | L mesio-temporal | L temporal atrophy (mesial & lateral) extending to insula & perisylvian area |
| 90 | 31-50 | L mesio-temporal | L MTS |
| 91 | 31-50 | R temporo-insular (posterior) | R temporal neocortex atrophy atrophy/agenesis of the R piriform |
| 92 | 18-30 | L mesio-temporal | L hemispheric atrophy, inferior frontal convexity polymicrogyria & MTS |
| 93 | 18-30 | B mesio-temporal | B MTS |
| 94 | 31-50 | R mesio-temporal | R MTS |
| 95 | 18-30 | L temporo-occipital | B mesial occipital ulegyria |

L: left, R: right, B: bilateral, FCD: focal cortical dysplasia, PNH: periventricular nodular, PCA: posterior cerebral artery, heterotopia, MTS: mesial temporal sclerosis, SMA: supplementary motor area.

### Supplementary Figure 1: Graphical User Interface

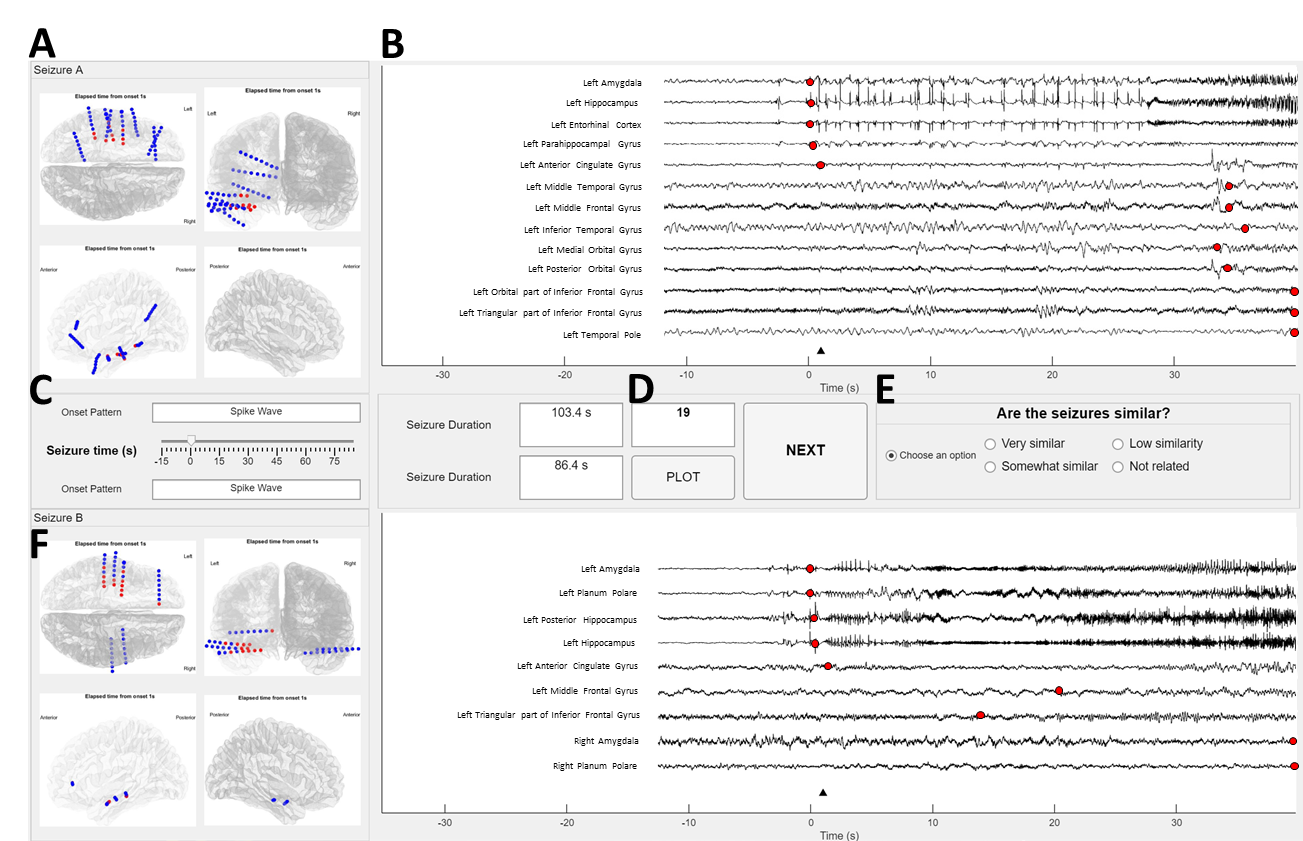

**Supplementary Figure 1: Graphical User Interface (GUI) for scoring seizure similarity**. (A) Axial, sagittal, and coronal views of the implantation scheme for seizure A. (B) Anatomical location of SEEG channels and corresponding segments with marked seizure onset (red) and end (blue). We illustrate 100 seconds of SEEG data (15 s preceding the onset of the seizure and 85 s succeeding the onset of the seizure) as assessed by experts. The SEEG shown in the figure is filtered between 0.3-100 Hz along with a notch filter to remove the powerline interference. The RED marker indicates the onset of the seizure in each channel, and the BLUE marker indicates the end of the seizure. The illustration shows the first ten regions and SEEG segments involved in the seizure. The white matter channels were removed. If a region has multiple channels implanted, we illustrate the channel which show the earliest seizure. There is an additional option to zoom in and out of the plot to see the SEEG patterns and the rest of the brain regions involved in the seizure. For seizure A (C) Seizure onset pattern, total duration, and seizure time slider for both the seizures. (D) Seizure pair index and navigation buttons (Plot, Next). (E) 4-level similarity score panel. (F) Panel for seizure pair B. The GUI description and code are available on https://github.com/Lab-Frauscher/Graphical-user-interface-for-seizure-similarity.

### Supplementary S1: Implementation of similarity features across seizure pairs

We investigated six similarity features across seizures. Euclidean distance was used to compute distance between two SEEG bipolar channels. If $\left( x_{i} {, y}_{i}, z_{i} \right)$ and $\left( x_{j} {, y}_{j}, z_{j} \right)$ are the Cartesian coordinates of two SEEG bipolar channels, *i* and *j*, the Euclidean distance between them is defined as:

$d_{i,j}=\sqrt{\left( x_{i}-x_{j} \right)^{2}+ \left( y_{i}-y_{j} \right)^{2}{+ \left( z_{i}-z_{j} \right)}^{2}}$	(1)

The maximum distance between two regions (centroid-to-centroid) across the MICCAI Atlas^23^ was 152 mm. To ensure consistency, all distance-based features were normalized using this maximum distance.

Seizure onset region: The onset region of a seizure is defined as the channels involved in the seizure within 500 milliseconds of the seizure onset. Let $\{C_{i}, i=1 to n\}$ and $\left\{ C_{j}, i=1 to m \right\}$ denote the set of onset channels corresponding to seizures A and B, respectively. The similarity between the onset regions of a pairs of seizures is calculated as:

$f_{1}=1-\frac{\bar{D}}{152}$, (2)

where

$\bar{D}= \frac{1}{N}\sum_{N} \{D_{A,B}, D_{B,A}\}$. (3)

and

$D_{A,B}=\left\{ \min\left\{ d_{C_{i},C_{j}},\forall j\in\left\{ 1,2,\ldots,m \right\} \right\},\forall i\in\left\{ 1,2,\ldots,n \right\} \right\}$. (4)

Seizure propagation region: The propagation region of a seizure is defined as the channels involved in the seizure between 500 and 5000 milliseconds of the of the seizure onset. We calculated the similarity between the propagation channels of a pairs of seizures ($f_{2}$) similar to the onset region.

Seizure duration: The seizure duration is defined as the time between the seizure onset and end. Let $t_{A}$ and $t_{B}$ denote the duration of seizures A and B, respectively. The similarity in duration between pairs of seizures is calculated as:

$f_{3}=1-\frac{\left| t_{A}-t_{B} \right|}{max\{t_{A},t_{B}\}}$. (5)

Seizure spread: Seizure spread is defined as the average propagated distance travelled by the seizure, starting from the onset and ending at 5000 milliseconds. Here, before computing the distance, we mapped the coordinates of the SEEG bipolar channels to the corresponding centroid of the brain regions according to the MICCAI atlas ($C\to R)$. Let $\{R_{i}, i=1 to n\}$denote the set of onset regions of the seizure and $\{R_{j}, j=1 to m\}$ denote the set of propagated regions within 5000 milliseconds. Seizure spread $p_{A}$ for seizure A is calculated as:

$p_{A}= \frac{\bar{D}}{152}$, (6)

where

$\bar{D}=\frac{1}{N}\sum_{N} \{D_{R_{i},R_{j}}\},$	(7)

and

$D_{R_{i},R_{j}}=\left\{ d_{R_{i},R_{j}},\forall j\in\left\{ 1,2,\ldots,m \right\} \right\},\forall i\in\left\{ 1,2,\ldots,n \right\}$	(8)

Let $p_{A}$ and $p_{B}$ denote the spread of seizures A and B, respectively. The similarity in spread between pairs of seizures is calculated as:

$f_{4}=1-\frac{\left| p_{A}-p_{B} \right|}{max\{p_{A},p_{B}\}}$. (9)

Seizure propagation speed: Seizure propagation speed is defined as the maximum propagated distance travelled by the seizure, starting from the onset region and ending at 5000 milliseconds divided by the duration. Similar to seizure spread, before computing the distance, we mapped the coordinates of the SEEG bipolar channels to the corresponding centroid of the brain regions according to the MICCAI atlas ($C\to R)$. Let $\{R_{i}, i=1 to n\}$denote the set of onset regions of the seizure, $\{R_{j}, j=1 to m\}$ denote the set of propagated regions within 5000 milliseconds, and $t_{5000}$ be the time taken from the onset to reach the final propagated region. Seizure speed $v_{A}$ for seizure A is calculated as:

$v_{A}= \frac{D_{max}}{t_{5000}}$, (10)

where

$D_{max}=\frac{max\{D_{R_{i},R_{j}}\}}{152}$	(11)

and

$D_{R_{i},R_{j}}=\left\{ d_{R_{i},R_{j}},\forall j\in\left\{ 1,2,\ldots,m \right\} \right\},\forall i\in\left\{ 1,2,\ldots,n \right\}$	(12)

Let $v_{A}$ and $v_{B}$ denote the speed of seizures A and B, respectively. The similarity in propagation speed between pairs of seizures is calculated as:

$f_{5}=1-\frac{\left| v_{A}-v_{B} \right|}{max\{v_{A},v_{B}\}}$. (13))

Seizure onset pattern: Seizure onset patterns are determined by the consensus of two epileptologists, according to the seven-pattern classification described by Perucca et al^25^. We defined a subjective correlation matrix (Supplementary Figure 2) for each pattern based on the opinion of two epileptologists and used these similarity values as a feature ($f_{6}$) for comparing pairs of seizures. The inclusion of this subjective correlation was necessary to acknowledge the presence of subjectivity in the characterization of certain seizure onset patterns by experts. In addition, it also facilitated the calculation of correlation between the onset pattern feature and expert labels. Omitting this subjective correlation would render the feature binary, either similar (1) or dissimilar (0).

### Supplementary Figure 2: Subjective correlation matrix for seizure onset patterns

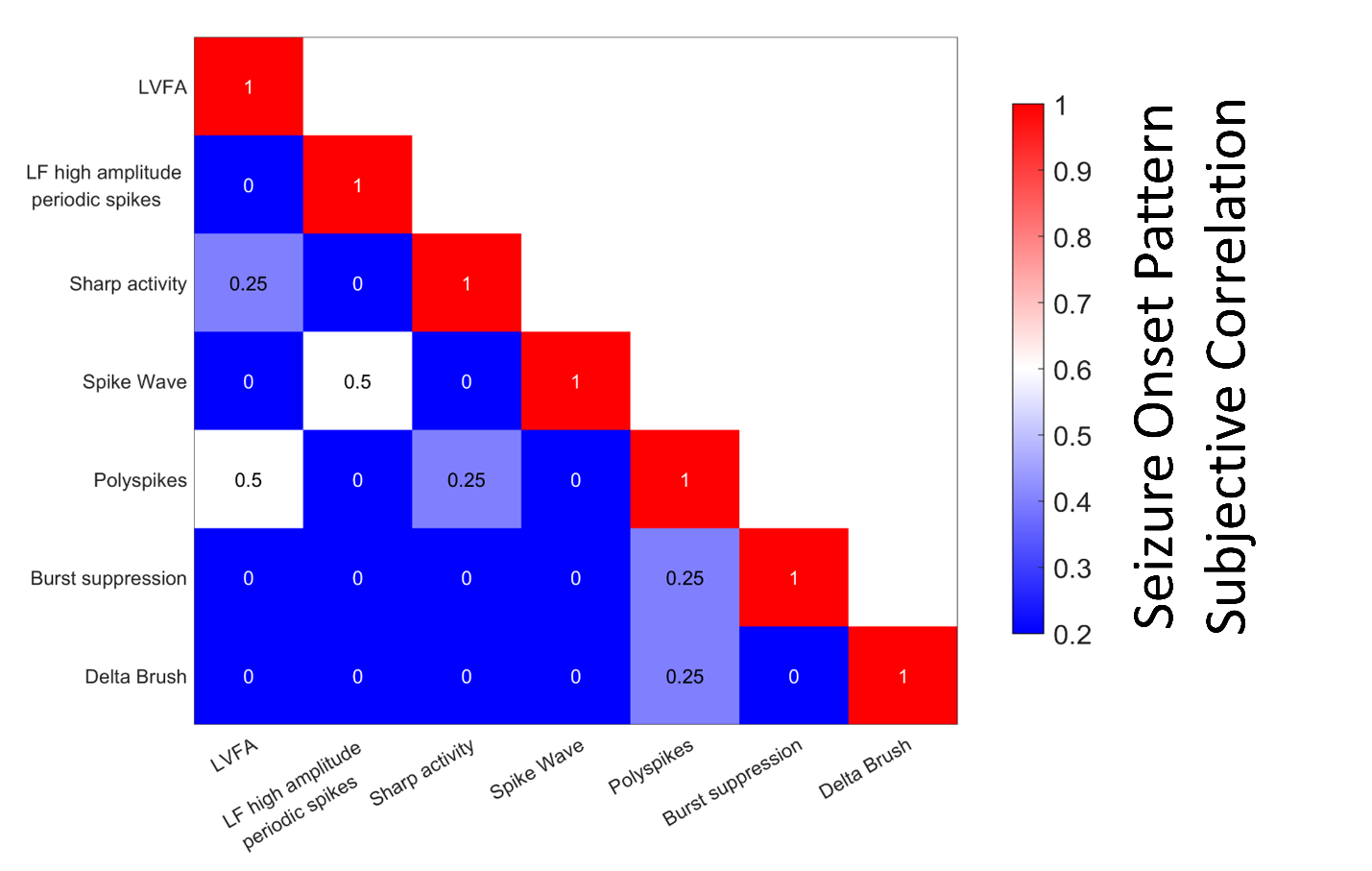

**Supplementary Figure 2: Subjective correlation matrix for seizure onset patterns.** We characterized the seizure onset patterns according to the seven-pattern classification described by Perucca et al. This correlation matrix was derived based on the opinion of epileptologists and we used these similarity values as features for comparing pairs of seizures. For, example, if seizure A shows an onset pattern ‘Polyspikes’ and seizure B shows an onset pattern of ‘Sharp activity’, the similarity value of 0.25 was used. This subjective correlation was utilized as a precaution to avoid ambiguity in determining seizure onset patterns as a standard definition of patterns in not available.

### Supplementary Figure 3: Feature correlation matrix

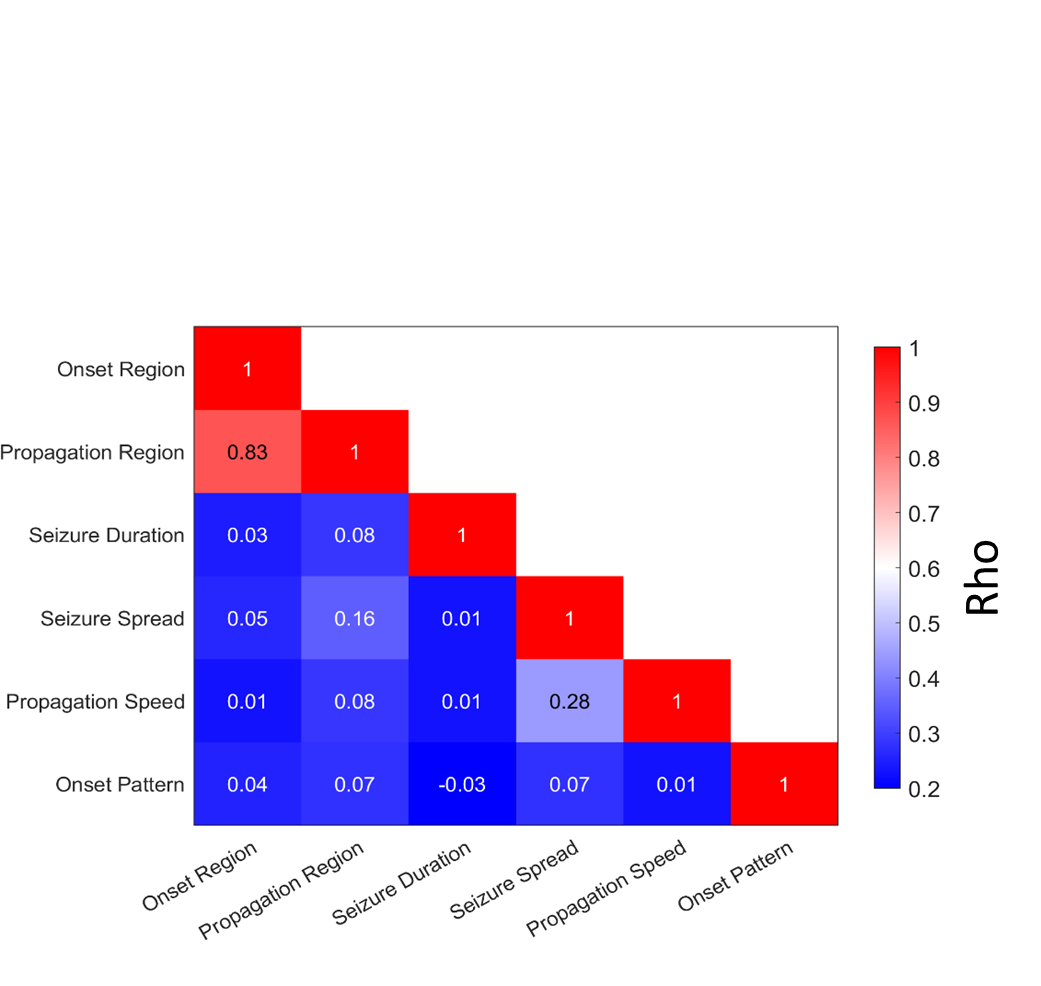

**Supplementary Figure 3: Feature correlation matrix.** The correlation between the six features were evaluated by computing the Spearman rho between feature values across all the possible pairs of seizures. A higher value of rho indicates a better correlation between the features. Out of the six features, the seizure onset region and propagation region were correlated.

### Supplementary S2: Survey document

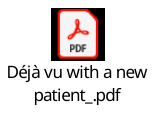
